# Wildfire smoke leaves a large five-year imprint on mortality in the United States

**DOI:** 10.64898/2026.10.15.26365428

**Authors:** Wenxiao Zhang, Hang Su, Ruijing Ni, Shu Tao, Jos Lelieveld, Yafang Cheng

**Author notes:** Correspondence to: Y.C. or H.S.

## Abstract

Wildfire smoke poses a growing health risk worldwide, yet the magnitude and temporal evolution of mortality after episodic exposure remain poorly resolved, limiting burden assessment. Using monthly county-level wildfire PM_2.5_ and mortality data from the United State, we exploit episodic exposure contrasts to deconvolve overlapping mortality responses and recover the impulse-response function. The excess mortality response peaks approximately three years after an exposure pulse and subsides by five years. We estimate 1.7 (95% uncertainty interval: 1.1–2.3) million smoke-associated deaths during 2011–2023, approximately ten times estimates using long-term ambient PM_2.5_ mortality relative risks. Circulatory and respiratory diseases contribute comparably and dominate this burden. Responses are markedly greater among adults ≥65 years; during the COVID-19 pandemic, population-wide cumulative responses are significantly higher over at least the first seven months after exposure. Reducing wildfire smoke exposure can thus yield substantial health benefits over multiple years, particularly when population vulnerability is heightened.

## Main text

Climate warming is intensifying wildfire danger in North America, a major global hotspot for extreme fire activity (*1, 2*). Wildfire smoke carries fine particulate matter (PM_2.5_; aerodynamic diameter ≤2.5 *μ*m) far beyond burned areas, posing a major public health threat and increasing mortality risk (*3*–*5*). The health effects of sustained exposure to anthropogenic PM_2.5_ are well established (*4, 6*–*8*). Wildfire smoke exposure, by contrast, is episodic, with strong seasonal and interannual variability (*3, 9*) (fig. S1). Yet for such episodic exposures, the cumulative health consequences of repeated exposure remain poorly understood, leaving an important uncertainty in assessing the health burden (*10, 11*).

A key unresolved question concerns the temporal response function linking an exposure pulse to subsequent mortality: how large is the response, how does it evolve, and when does it subside? If the response extends beyond the lag window used in an assessment, delayed deaths may be missed. On the other hand, long averaging windows can blur exposure pulses, making delayed mortality effects difficult to identify. Recurrent smoke exposures further complicate identification of this temporal response: when new exposures occur before earlier responses have subsided, mortality in a given period can reflect the overlapping effects of multiple prior exposures.

To address this challenge, we build on a deconvolution-based distributed-lag framework (*12*–*14*) and adapt it to the temporal structure of recurrent wildfire PM_2.5_ exposure. Using monthly county-level wildfire PM_2.5_ concentrations (*3, 9*) and mortality records (*15*) across the contiguous United States (US) from 2006 to 2023 (fig. S1), we exploit the episodic feature of wildfire smoke exposure to disentangle overlapping effects and recover the mortality impulse-response function associated with an individual exposure episode. We then use the estimated contemporaneous and delayed responses to quantify the cumulative mortality burden associated with wildfire PM_2.5_ exposure.

To isolate the effect of wildfire PM_2.5_, our analysis non-parametrically accounts for several classes of confounders. We include fixed effects that absorb time-invariant differences across counties (for example, geographic characteristics and industrial structure), national month-of-sample effects that capture coherent temporal variation across all counties (such as broad socioeconomic or policy changes), and county-specific temporal components that flexibly represent seasonal cycles, interannual variability, and other nonlinear trends (including local holidays, policy interventions introduced in specific years, and gradual improvements in healthcare systems). In addition, we incorporate ambient temperature parametrically, which has been shown to exert nonlinear effects on mortality (*16*) and to influence wildfire activity (*17*). Wildfire PM_2.5_ exposure is modeled as a sequence of monthly impulses at the county level, and mortality outcomes reflect the cumulative contributions of contemporaneous and prior exposures. We recover the characteristic impulse-response function by estimating a set of distributed lag coefficients that, when applied jointly across all exposure episodes, best reproduce the observed mortality time series (detailed in Materials and Methods M2.1–M2.4; figs. S2 to S3).

### Impulse-response of mortality to wildfire PM_2.5_ exposure

Figure 1A shows the recovered impulse-response function, which describes how the excess mortality rate evolves over time after a wildfire PM_2.5_ exposure episode. Monthly mortality rates exhibit a slight increase immediately following exposure, then rise gradually from baseline to a delayed peak of 0.0035 (95% confidence interval, CI: 0.0025–0.0045) deaths per 1,000 population per 1 *μ*g m^−3^ increment in wildfire PM_2.5_ at approximately 30–36 months (~three years). Thereafter, the excess mortality rate declines, returning towards baseline at approximately 54–60 months. These results indicate that a single wildfire PM_2.5_ exposure episode is associated with an elevated mortality rate that persists for about five years. The corresponding cumulative effect, obtained by summing monthly responses, increases gradually and then levels off, reaching a total increase of 0.078 (95% CI: 0.050–0.11; P < 0.01) deaths per 1,000 population per 1 *μ*g m^−3^ increment in wildfire PM_2.5_ over 60 months following exposure (orange solid curve in Fig. 1B). Note that the estimates presented here are from our “*main model*”, fitted using mortality data from 2011 to 2019 and exposure histories from 2006 to 2019 (Materials and Methods M2.1). The potential influence of the COVID-19 pandemic is explored separately using mortality data from 2020 to 2023 and addressed in the Discussion.

**Fig. 1.**
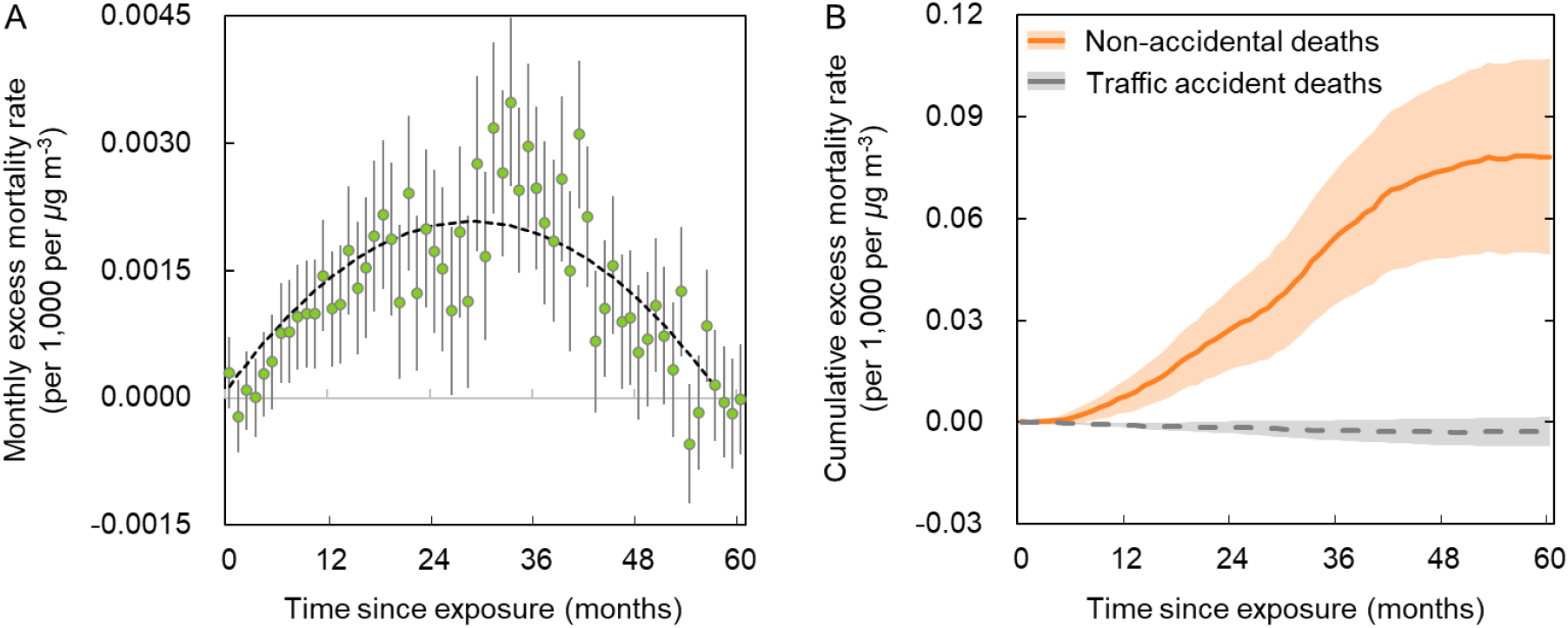
Excess mortality following a wildfire PM_2.5_ exposure episode. **(A)** Impulse-response function for non-accidental mortality associated with wildfire PM_2.5_, showing monthly excess mortality rates in the following 60 months after exposure. The black dashed line shows a quadratic fit to the monthly response estimates. Error bars show 95% confidence intervals. **(B)** Wildfire PM_2.5_-associated cumulative excess mortality rates for non-accidental deaths and traffic accident deaths (a negative outcome control), obtained by summing monthly responses up to 60 months after exposure. Shaded bands show 95% confidence intervals (accounting for covariances). Both panels show responses associated with a 1 *μ*g m^−3^ increment in wildfire PM_2.5_ exposure in a given month, expressed per 1,000 population. Estimates are from our main model using mortality data from 2011 to 2019 and exposure histories from 2006 to 2019.

Our model optimally balances parsimony and explanatory power, adequately capturing historical variation in non-accidental mortality rates at both national and county levels, with strong predictive skill (R^2^ = 0.924; figs. S2 to S3). To assess robustness, we follow Young and Hsiang (*12*) and conduct a suite of placebo tests including associations with future wildfire PM_2.5_ exposure and multiple reshuffling schemes (unconditional fully, within-county, within-time, and across-county randomization) (Materials and Methods M2.3). Across all placebo tests, estimated effects are centered near zero and fail to reproduce the patterns observed in the main model (figs. S4 to S5). Our model indicates no spurious associations between mortality rates and exposure in these tests, whereas alternative models that omit any of the major confounders fail at least one of these falsification tests or residual diagnostics (Supplementary Text S3; figs. S6 to S7). As an additional negative outcome control, applying the same deconvolution to traffic-accident mortality rates yields cumulative effects centered around baseline (grey dashed curve in Fig. 1B). Together, these diagnostics suggest that the estimated wildfire PM_2.5_– mortality associations are unlikely to be driven by random noise or by the major confounding structures addressed by our design.

### Cumulative mortality burden

We apply the impulse-response function recovered from our main model to each monthly exposure pulse through 2023 to reconstruct the associated mortality trajectory over the following 60 months. Monthly wildfire PM_2.5_-associated deaths are then calculated by summing contributions from current and prior exposures to deaths occurring in that month. These estimates account for changes in county populations over time (Materials and Methods M2.5). The superposition of overlapping responses produces a gradual temporal pattern in monthly mortality (black curve in Fig. 2A), rather than the pronounced seasonal spikes observed in monthly wildfire PM_2.5_ exposure (Fig. 2B). For example, annual smoke-associated deaths are estimated at 91,000 (95% uncertainty intervals, UI: 61,000– 122,000) in 2018 and rise to 289,000 (95% UI: 192,000–394,000) in 2023 (Fig. 2A). The elevated mortality in 2023 reflects both high wildfire smoke exposure in that year and the cumulative impact of successive intense fire seasons in preceding years.

**Fig. 2.**
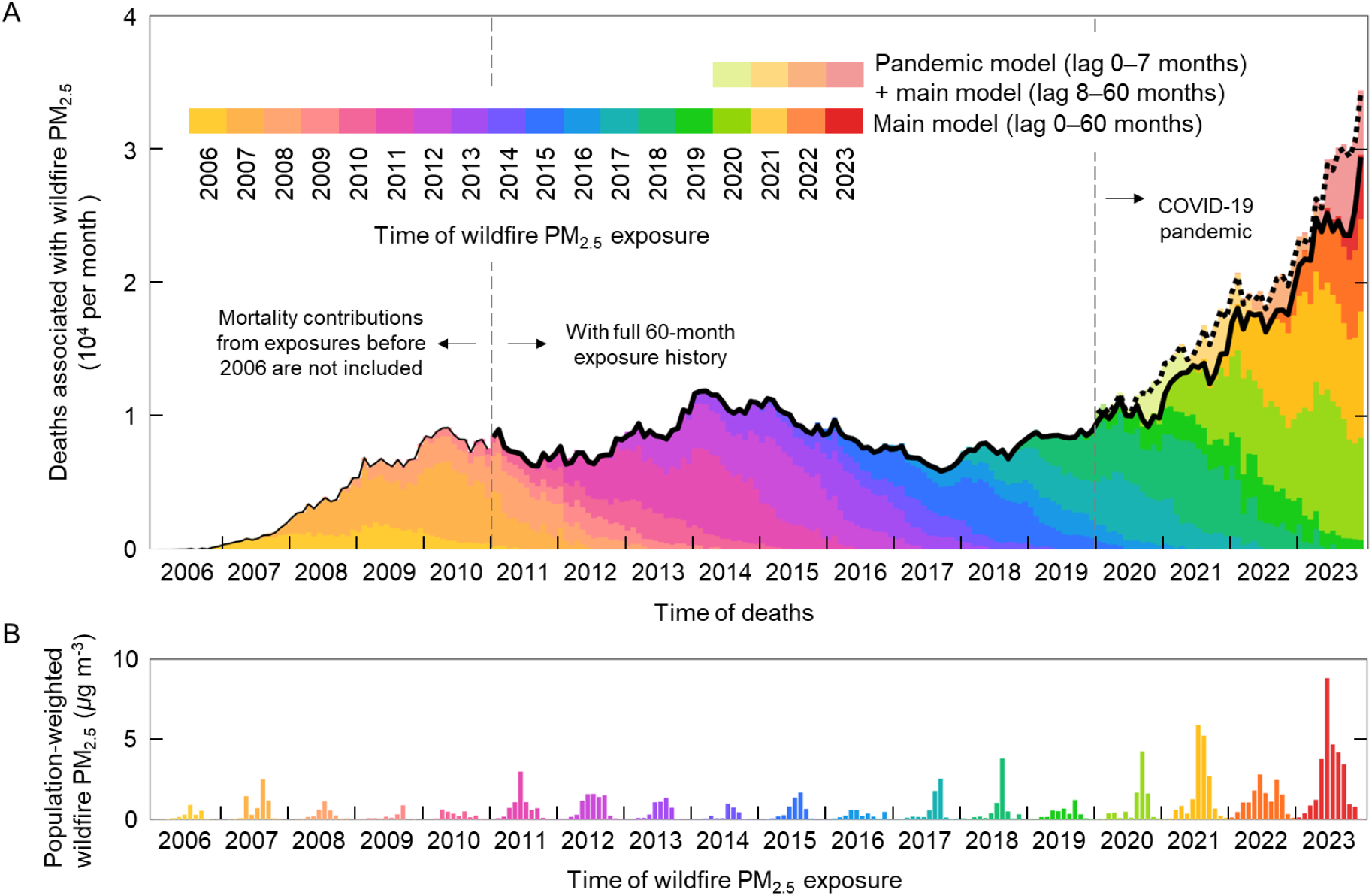
Temporal distribution of wildfire PM_2.5_ exposure and associated deaths in the contiguous United States. **(A)** Non-accidental deaths associated with wildfire PM_2.5_ exposure during 2006–2023. Main model estimates are shown by saturated colors and solid black lines. Muted colors and a thick dashed black line show estimates for deaths occurring during the COVID-19 pandemic period (2020–2023), combining coefficients from the pandemic model at lag 0–7 months with those from the main model at lag 8–60 months. Black lines are thin before 2011 because mortality contributions from exposures before 2006 are not included, and thick from 2011 onward when the full 60-month exposure history is available. The colors of the stacks identify the years of exposure; their positions along the x-axis indicate when the associated deaths occur. **(B)** Monthly population-weighted mean wildfire PM_2.5_ concentrations across counties in the contiguous US during 2006–2023.

During 2011–2023, an estimated 1.7 (95% UI: 1.1–2.3) million deaths in the contiguous US are associated with wildfire PM_2.5_ exposure, averaging 130,000 (95% UI: 86,000–176,000) deaths per year. Since exposure records begin in 2006, we report mortality burdens from 2011 onward to retain a complete 60-month exposure history (thick black curve in Fig. 2A). The estimated annual burden far exceeds officially reported direct wildfire deaths—primarily from burns, trauma, and other acute injuries—which typically number in the tens to around a hundred annually, even during extreme fire seasons (*18*) (fig. S8). Our estimated burden represents 5.2% (95% UI: 3.4%–7.1%) of total deaths during this period—substantially higher than the 0.27% reported for acute wildfire PM_2.5_-associated mortality in North America during 2000–2015 reported in a previous global time series study using a 3-day moving average of exposure (lag 0–2 days) (*5*).

For comparison, we estimated wildfire smoke-associated deaths during 2011–2023 by applying published mortality relative risk from a global meta-analysis and a US cohort study of long-term exposure to total ambient PM_2.5_ (*7, 8*) (Supplementary Text S4). This approach yields 12,000–18,000 deaths per year—approximately 9–14% of the 130,000 deaths estimated for this period using our wildfire-specific 60-month impulse-response function. These findings suggest that wildfire smoke represents a fundamentally different class of climate-related health hazard: episodic wildfire smoke exposure can leave a mortality imprint lasting for years and impose a large cumulative health burden not previously captured by conventional approaches.

### Spatial and temporal patterns in mortality burden

Figure 3A shows the spatial distribution of mean wildfire PM_2.5_-associated mortality rates during 2011-2023. These rates reflect the lagged contributions of smoke exposure over the preceding 60 months and broadly follow the distribution of mean wildfire PM_2.5_ concentration (fig. S1). Rates are highest in the northwestern US and in northern and central California, with more moderate elevations across parts of the Great Plains. Absolute death counts (Fig. 3B), which reflect both mortality rates and county populations, show a broader geographic distribution, with greater prominence in southern California and in populous counties across Texas and the eastern US, including Florida. Large populations can therefore translate relatively low mortality rates into substantial death counts.

**Fig. 3.**
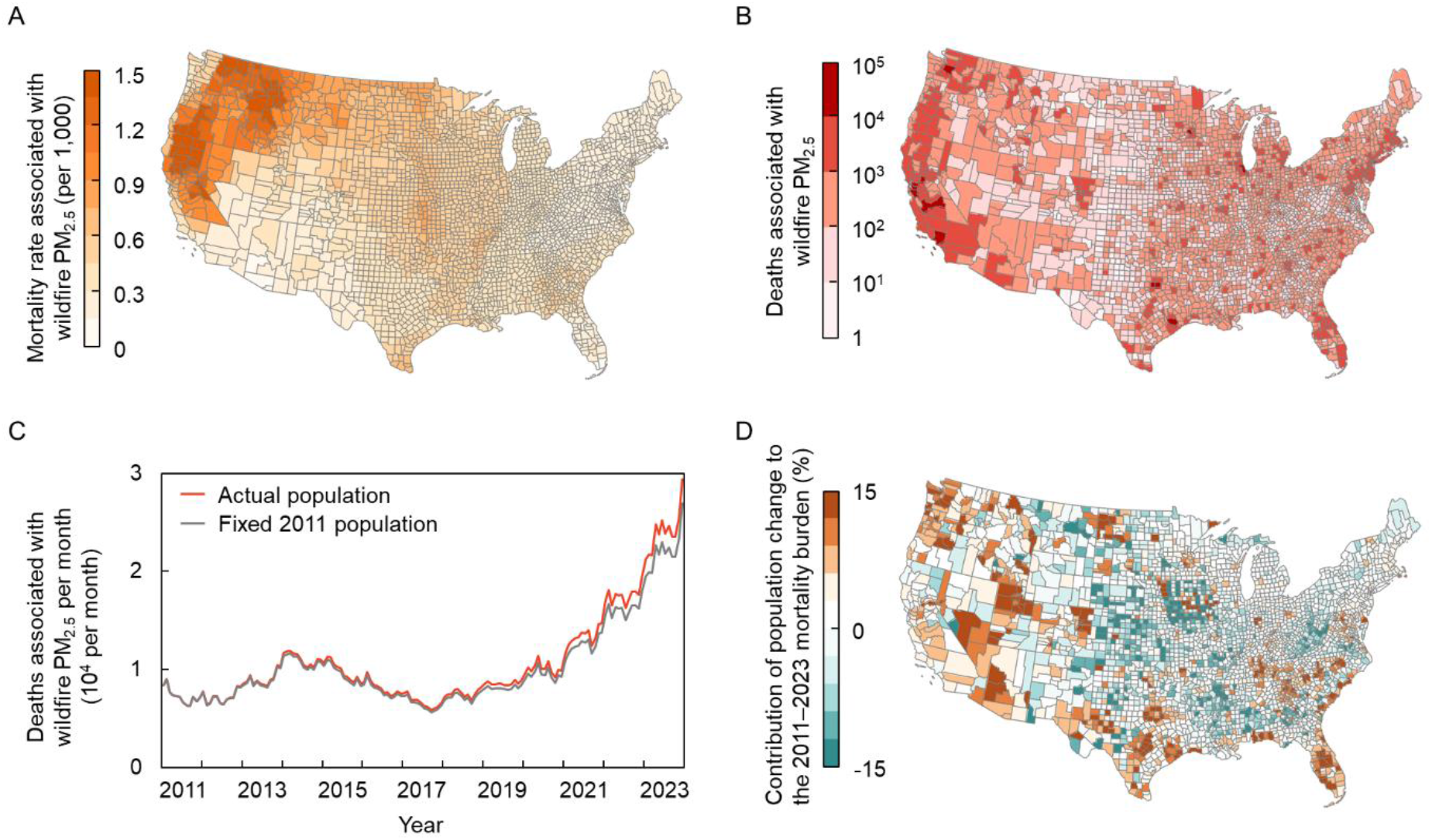
Spatial patterns of wildfire PM_2.5_-associated non-accidental deaths in the contiguous United States during 2011–2023. (**A**) Average annual smoke-associated mortality rate. (**B**) Total smoke-associated deaths during 2011–2023. (**C**) Monthly smoke-associated deaths estimated using actual county populations and county populations held fixed at their 2011 levels. (**D**) Percentage contribution of population change to the smoke-associated mortality burden during 2011–2023. This contribution is calculated as the difference between deaths estimated using actual and fixed populations, expressed as a percentage of the estimated deaths under actual populations.

From 2011 to 2023, estimated annual wildfire PM_2.5_-associated deaths in the contiguous US increase approximately 3.3-fold, from 87,000 (95% UI: 62,000–119,000) to 289,000 (95% UI: 192,000– 394,000), with most of the net increase occurring after 2018. Most of this increase persists when county populations are held fixed at their 2011 levels (Fig. 3C), indicating that changes in smoke exposure and their mortality effects are the primary drivers. Estimated wildfire PM_2.5_-associated annual mortality rates also rise from about 0.28 to 0.86 deaths per 1,000 population over this period. Meanwhile, the US population grew from 307 million to 335 million, with many western counties that were already populous in 2011 also experiencing growth (fig. S9). Relative to the fixed-population counterfactual, population growth and redistribution account for 5.0% of the total estimated mortality burden during 2011–2023. Population change increases the estimated burden in many western and southeastern counties but reduces it across parts of the central US (Fig. 3D). For example, population change accounts for 10.6% and 10.2% of the estimated mortality burden in Washington and Texas, respectively, despite considerably lower mean smoke concentrations in Texas (fig. S10). Population growth and redistribution can therefore amplify mortality burdens in both highly exposed regions and those with lower smoke exposure.

### Demographic heterogeneity in vulnerability

We further estimate the mortality response of wildfire PM_2.5_ exposure by age, sex, and race, to assess variations across population groups (Materials and Methods M2.1). Among these demographic dimensions examined, age shows the largest contrast in cumulative mortality responses to wildfire PM_2.5_ exposure. A 1 *μ*g m^−3^ increase in exposure in a given month is associated with 0.46 (95% CI: 0.30–0.61; P < 0.01) additional deaths per 1,000 adults aged ≥65 years over the subsequent 60 months, compared with 0.021 (95% CI: 0.0047–0.038; P = 0.012) deaths per 1,000 people aged <65 years—a roughly 20-fold contrast (Fig. 4A). This pronounced age difference is consistent with a previous fixed-effects analysis using 12-month moving-average wildfire PM_2.5_ exposure, in which mortality-rate responses among older adults are approximately 5.36–22 times those among younger individuals across concentration categories (*19*). It also aligns with evidence of greater PM_2.5_-associated cardiorespiratory hospitalization risks among older adults (*20*). To compare mortality burdens across study designs, we apply the published wildfire smoke–mortality association from a southern California cohort of adults aged ≥60 years to the region’s population aged ≥65 years (*21*) (Supplementary Text S4). This yields approximately 37,000 smoke-associated deaths during 2011–2019—of the same order as our cumulative estimate of 22,000 deaths for the same population and period. Protecting older adults is thus especially critical amid ongoing population aging (*22*).

**Fig. 4.**
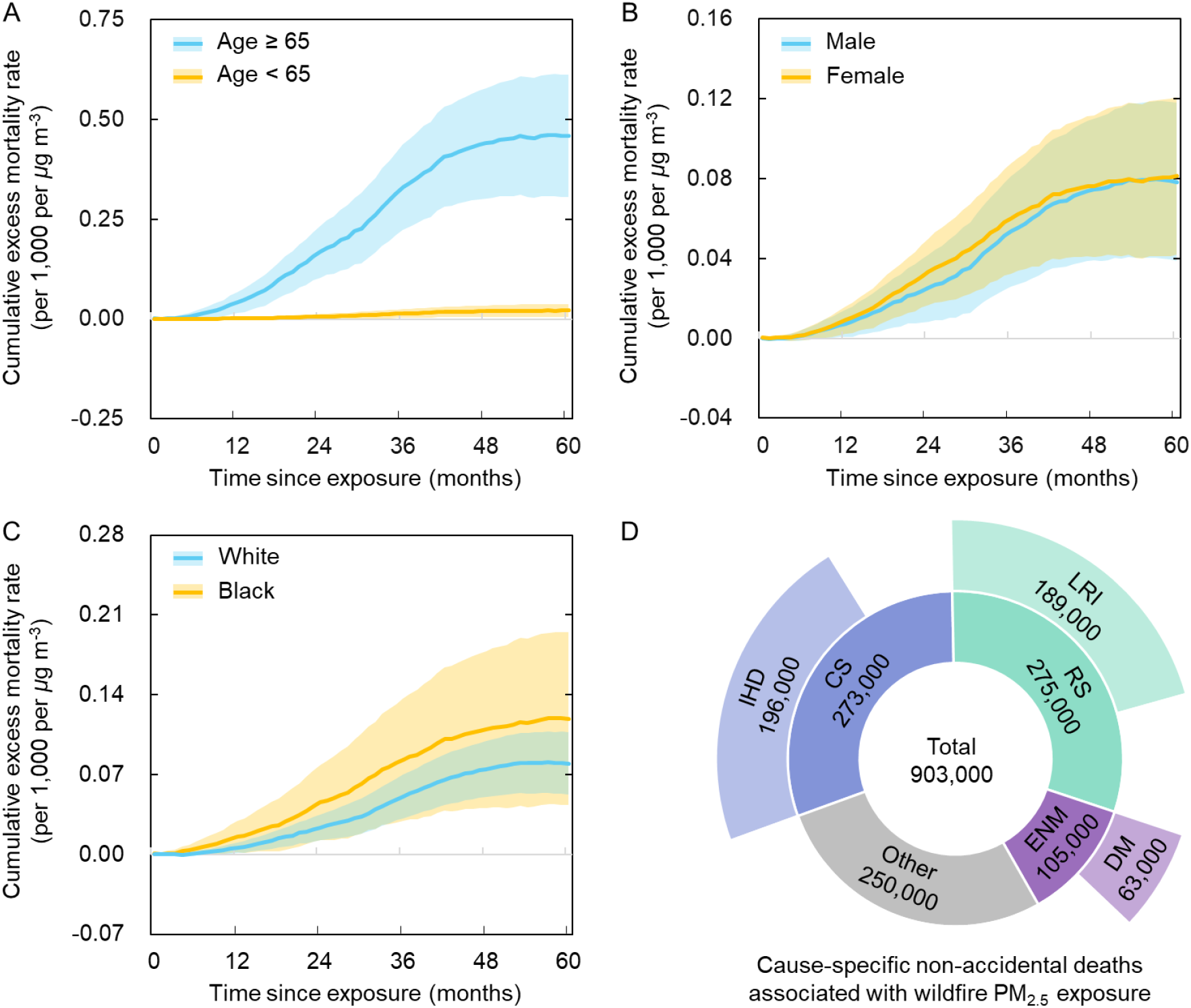
Subpopulation and cause-specific non-accidental mortality associated with wildfire PM_2.5_ exposure. Cumulative excess mortality rates across population subgroups by age **(A)**, sex **(B)**, and race **(C)**, obtained by summing monthly responses up to 60 months after exposure. Estimates correspond to a 1 *μ*g m^−3^ increment in wildfire PM_2.5_ exposure in a given month and are expressed per 1,000 population within each subgroup. Shaded bands show 95% confidence intervals (accounting for covariances). **(D)** Contributions of different causes of death to the total smoke-associated non-accidental mortality burden during 2011–2019. Ischemic heart disease (IHD), lower respiratory infections (LRI), and diabetes mellitus (DM) are shown as subcategories of diseases of the circulatory system (CS), diseases of the respiratory system (RS), and endocrine, nutritional, and metabolic diseases (ENM), respectively.

Cumulative mortality-rate responses are broadly similar between females and males. Estimates are higher for Black than for White populations, with greater uncertainty among Black populations partly reflecting sparse county-level death counts and frequent zero observations (Fig. 4A). Based on these responses, the estimated share of non-accidental deaths associated with wildfire PM_2.5_ is higher among Black than White populations (7.2% versus 3.9%), partly reflecting lower baseline non-accidental mortality rates among Black populations in our data. These sex- and race-specific response patterns align with findings from a nationwide cohort of adults aged ≥65 years, which reports no significant sex differences in smoke–mortality associations but higher hazard ratios among Black than White participants (*23*). It suggests that while age dominates the observed vulnerability patterns, racial differences may be more apparent in relative mortality burden than in absolute effect sizes.

### Disease-specific mortality burden

Our cause-specific analyses identify circulatory (CS), respiratory (RS), and endocrine, nutritional, and metabolic (ENM) diseases as major contributors to the persistent mortality burden (Materials and Method M2.1). A 1 *μ*g m^−3^ increase in wildfire PM_2.5_ exposure in a given month is associated with 0.024 (95% CI: 0.006–0.041; P < 0.01), 0.024 (95% CI: 0.014–0.034; P < 0.01), and 0.009 (95% CI: 0.003– 0.016; P < 0.01) additional deaths per 1,000 population, respectively, accumulated over the 60 months following exposure (fig. S11). For CS and ENM, first-year cumulative responses are comparable in magnitude to estimates from a study using 12-month moving-average smoke exposure (*19*) (Supplementary Text S5). That study, however, finds no consistent positive association with respiratory mortality. We show that cumulative responses for all three disease categories continue to rise well beyond the first year, indicating that assessments confined to this initial period would miss substantial delayed mortality.

At the population level, wildfire smoke is associated with an estimated 273,000 circulatory, 275,000 respiratory, and 105,000 ENM deaths during 2011–2019, together accounting for about 72.3% of the total estimated smoke-associated mortality burden (Fig. 4D). Although circulatory and respiratory diseases contribute similar numbers of deaths, their relative burdens differ markedly: smoke-associated deaths represent 11.8% of all respiratory deaths, compared with 3.7% of circulatory deaths (fig. S12). This reflects the higher baseline mortality from circulatory diseases and underscores the substantial contribution of wildfire smoke to respiratory mortality.

Within these broad categories, ischemic heart disease (IHD; 196,000 deaths) and lower respiratory infections (LRI; 189,000 deaths) each account for approximately 70% of their respective circulatory and respiratory burdens (Fig. 4D). Their prominence aligns with evidence linking long-term ambient PM_2.5_ exposure to mortality from these causes (*4, 8*). Wildfire-specific evidence also links two-year average wildfire PM_2.5_ exposure to increased hospitalizations for IHD and pneumonia across 20 US states (*24*). Mechanistic studies have also reported impaired macrophage phagocytosis following exposure to bushfire smoke extracts and changes in circulating inflammatory and coagulation markers following controlled wood-smoke exposure (*25, 26*). The ENM burden is dominated by diabetes (DM), which accounts for approximately 63,000 deaths, or 60% of this category (Fig. 4D). Long-term ambient PM_2.5_ exposure has been associated with diabetes mortality in a US cohort (*27*), while experiments in mice fed a high-fat diet show that concentrated ambient PM_2.5_ exacerbates adipose inflammation and insulin resistance (*28*). Furthermore, combustion-related ultrafine particles can reach the alveoli and enter the systemic circulation, affecting distant organs and potentially increasing the risk of metabolic disease (*29, 30*). Together, the concentration of deaths in cardiopulmonary and metabolic diseases highlights priority for wildfire smoke-related health protection.

### Discussion and implications

The episodic nature of wildfire PM_2.5_ provides a temporal anchor for understanding how mortality risk unfolds and evolves after exposure, a dimension not captured by static exposure–response relationships or a predefined lag window alone. In a distributed-lag framework, mortality in a given period reflects overlapping contributions from current and past exposures. The recovered impulse-response function disentangles these contributions and provides several advantages. First, it reveals when excess mortality emerges, peaks, and subsides, identifying periods of elevated risk that may warrant closer monitoring and timely intervention. Second, the timing and shape of the overall mortality response may provide mechanistic clues by distinguishing delayed or sustained health consequences from acute effects. Third, resolving how mortality unfolds after an exposure episode can improve estimates of smoke-associated mortality by reducing reliance on predefined lag windows and the risk of omitting delayed effects. Finally, differences among cause-specific impulse-response functions can provide insights into the disease processes and potential mechanisms linking exposure to excess deaths.

Recovering this temporal function requires sufficient temporal contrast among exposures, which the episodic nature of wildfire smoke can provide. Wildfire episodes repeatedly perturb the system through exposure pulses that vary markedly in timing and intensity, providing information analogous to repeated excitation in a dynamic system. By contrast, slowly varying or persistently elevated exposure can produce strong correlations among lagged exposure values, making it difficult to disentangle the mortality contributions of exposures at different times, particularly in the presence of uncertainty in input data and other sources of noise. To examine how exposure temporal structure affects response recovery, we constructed two synthetic PM_2.5_ exposure series with the same underlying temporal exposure-response function but different temporal structures. One retained the episodic pattern of observed wildfire PM_2.5_, whereas the other resembled the smoother temporal variation of non-wildfire ambient PM_2.5_. At low noise levels, the underlying response could be recovered from both series. As noise increased, however, the recovery from the smoother series became progressively distorted, whereas recovery from the episodic series remained substantially more faithful to the underlying function (Supplementary Text S6). These simulations demonstrate that exploiting the episodic nature of wildfire smoke exposure can improve recovery of the temporal response under noisy conditions, highlighting a fundamental identifiability advantage conferred by the temporal contrast inherent in episodic exposure.

Temporally averaged exposure metrics blur the episodic structure of wildfire smoke exposure: brief, intense smoke episodes and longer periods of less intense exposure can yield the same mean concentration (*11*). This loss of temporal contrast can increase uncertainty in lagged estimates and mask biologically meaningful temporal structure, limiting identification of prolonged responses. Comparisons with studies using moving-average or annual exposure metrics further underscore the importance of retaining this temporal information. As shown in fig. S13, over matched one-to two-year exposure windows, the findings of Ma et al. (*19*) and Qiu et al. (*1*) are broadly consistent with ours in identifying increased mortality. However, as their averaging or lag windows extend to three years or more, estimates attenuate toward the null or become unstable, preventing reliable characterization of the response dynamics—when excess mortality emerges, peaks, and subsides—and leaving the full cumulative burden unresolved (Supplementary Text S5).

By resolving the temporal exposure–response function, our framework transforms the conventional concept of “lag” into a measurable dynamic trajectory of post-exposure risk. In this sense, lag carries statistical as well as biological information: differences in the timing, shape, and persistence of mortality responses may reflect pathophysiological processes unfolding on different timescales. Uncertainty in wildfire PM_2.5_ exposure estimates and mortality data, together with limited record length and spatial and temporal resolution, constrains our ability to resolve finer temporal features and to interpret their potential biological significance. However, longer records of high-quality data with finer spatial and temporal resolution—including weekly or daily observations—are expected to improve exposure assignment and linkage to disease-specific mortality records, better resolve the timing and shape of the response, and reduce uncertainty. Linking such data to longitudinal clinical records could help distinguish delayed mortality from acute effects and inform hypotheses about how wildfire smoke exposure may influence disease progression over time.

Our wildfire-specific 60-month impulse-response function yields a mortality burden approximately ten times that estimated using conventional mortality relative risks of long-term exposure to ambient PM_2.5_ during 2011–2023. This suggests that transferring ambient PM_2.5_ response functions to wildfire smoke can substantially underestimate its mortality burden. Recent nationwide cohort evidence likewise reports significantly stronger all-cause mortality associations per unit concentration for smoke than non-smoke PM_2.5_ among older US adults. The same study reports particularly strong associations with smoke-derived carbonaceous components, providing epidemiological support for particle composition as a contributor to source-specific health effects (*23*). These findings complement evidence of stronger respiratory and multiyear cardiovascular hospitalization associations for wildfire than non-wildfire PM_2.5_ (*31, 32*). Experimental comparisons also show greater oxidative-stress and xenobiotic responses to wildfire than urban particles (*33*). Combustion of structures and household materials can add hazardous organic compounds and metals to smoke (*34*). A recent study of adults assessed 6–18 months after the 2023 Maui wildland–urban interface fires found elevated urinary metal concentrations relative to US reference levels, with higher metal-mixture burdens associated with impaired lung function (*35*). Particularly harmful particle composition and co-emitted pollutants in wildfire smoke may contribute to these strong health effects.

Population vulnerability may further amplify these impacts. We explore this possibility in the context of the COVID-19 pandemic by comparing mortality responses estimated from 2020–2023 mortality data (“*pandemic model*”) with the pre-pandemic main model response (Materials and Methods M2.1). The pandemic model yields a significantly greater cumulative mortality response over lag 0–7 months (fig. S14). Cumulative responses remain higher at longer lags, although the differences are not statistically significant, which may partly reflect greater uncertainty associated with the shorter pandemic-period mortality record. This comparison uses non-accidental deaths, excluding those recorded as COVID-19 deaths, and accounts for temporal trends in both periods. To assess the implications of the early enhancement, we replace the coefficients for lag 0–7 months with pandemic model estimates when calculating deaths occurring during 2020–2023, while retaining the main model coefficients at longer lags (lag 8–60 months). This further increases the estimated burden by 13.5% during 2020–2023 and by 6.3% over 2011–2023 (Fig. 2A). Multisystem sequelae following SARS-CoV-2 infection may have contributed to heightened vulnerability during the pandemic (*36*–*38*). Together with the substantially larger responses among adults aged ≥65 years, these findings underscore the importance of population vulnerability.

Our study demonstrates the need for future health assessments to account for the magnitude, persistence and temporal evolution of wildfire-specific mortality effects, together with changes in population vulnerability. Reducing smoke exposure can yield substantially greater health benefits than conventional estimates imply, with gains that extend for multiple years beyond the fire season.

## Data Availability

The data sources, models, and model configurations underlying the conclusions are described in the main text.

## Acknowledgments

This study is supported by the Max Planck Society (MPG). Yafang Cheng, Hang Su and Wenxiao Zhang acknowledge the support of Max Planck-CAS-IITM-UHEL Center for “Unraveling the Nexus of Air Pollution, Extreme Weather, and Monsoon in a Warming Climate: Pathways to Global Solutions” and Max Planck-Caltech-Carnegie-Columbia Center for “Land-surface Mediated Earth System Feedbacks – AI Enabled Understanding, Modeling and Prediction”.

## Author contributions

Conceptualization: YC

Methodology: WZ, YC, HS, RN

Investigation: WZ, YC, HS, RN

Interpreted results: WZ, YC, HS

Discussed results: RN, JL, ST

Visualization: WZ, YC

Funding acquisition: YC, HS

Project administration: YC, HS

Writing – original draft: YC, HS, WZ

Writing – review & editing: RN, JL, ST

## Competing interests

The authors declare that they have no competing interests.

## Data and code availability

The data sources, models, and model configurations underlying the conclusions are described in the main text and Materials and Methods. The model and analysis code, together with the input and output files, will be made publicly available upon acceptance of the manuscript.

## Notes

### Competing Interest Statement

The authors have declared no competing interest.

### Author Declarations

Month-county level mortality data in the contiguous United States (US) from 2006 to 2023 were collected from the CDC (Centers for Disease Control and Prevention) WONDER (Wide-ranging Online Data for Epidemiologic Research) online database. (https://wonder.cdc.gov/)

